# Plasma proteomic evidence for increased Alzheimer’s disease-related brain pathology after SARS-CoV-2 infection

**DOI:** 10.1101/2024.02.02.24302132

**Authors:** Eugene P Duff, Henrik Zetterberg, Amanda Heslegrave, Abbas Dehghan, Paul Elliot, Naomi Allen, Heiko Runz, Rhiannon Laban, Elena Veleva, Christopher D Whelan, Benjamin B Sun, Paul M Matthews

## Abstract

Prior studies have suggested that systemic viral infections may increase risks of dementia. Whether this holds true for SARS-CoV-2 virus infections remains uncertain but is of great consequence for predicting future dementia rates. We examined this by comparing changes in plasma biomarkers in UK Biobank participants before and after serology confirmed SARS-CoV-2 infections. We discovered biomarker changes associated with increased AD risk within this population. SARS-CoV-2 infection was associated with reduced plasma Aβ42:Aβ40 concentration ratios, and in more vulnerable participants, lower plasma Aβ42 and higher plasma pTau-181. These biomarker changes, which have been associated with brain beta-amyloid accumulation in prodromal AD, were associated here with increased brain imaging signatures of AD, poorer cognitive scores, and worse assessments of overall health. Changes were greater in participants who had been hospitalised with COVID-19 or had previously reported hypertension. Our data provide evidence for the hypothesis that SARS-CoV-2 can be associated with accelerating brain pathology related to prodromal AD.

## Introduction

Exposures to infectious diseases in early life and adulthood have been linked to increased risk for neurodegenerative and other systemic disease ^1–4^. Viral encephalitis or meningitis, influenza, pneumonia and viral intestinal infections all may increase risks of Alzheimer’s disease (AD) ^1^, Vaccinations to prevent these diseases may also have protective effects ^5^. A variety of central and peripheral mechanisms could underly these effects, including tissue injury during the acute phase of infections or chronic secondary inflammatory processes^6–8^. However, uncertainty remains. The epidemiological observations are confounded by heterogeneity of infections, and mechanistic hypotheses have been limited by the lack of large scale, harmonised, prospective observations in human populations that allow common factors that predispose both to infection and to dementia to be discriminated from any potential causal relationships of infection to dementia.

The recent COVID-19 pandemic provided a common viral exposure to large populations over well-defined periods. SARS-CoV-2 initiates a systemic inflammatory response, which persists in many patients beyond the acute phase and increases susceptibility to several systemic diseases. Although the virus is not neurotrophic ^9–11^, impairment of cognition, and brain structural changes have been reported as sequelae of SARS-CoV-2 infections, even amongst people who did not require hospitalisation or experience long-COVID^10,12^. Initial evidence for raised dementia rates in vulnerable populations following serious COVID have been reported^13^. Preclinical studies have identified potential mechanisms for increased brain inflammatory pathology with SARS-CoV-2 that could account for these observations^7,14,15^. These reports suggested to us that the COVID pandemic could provide a “natural experiment” for testing whether infection-associated systemic infection can initiate or potentiate brain pathology associated with AD. The urgency of the question seemed clear: the global scale of the pandemic and aging populations make a better understanding of the potential impact of COVID-19 and other infections on the prevalence of future dementias a public health priority^10,16^.

Recently developed high-throughput plasma proteomic biomarkers that provide biologically-based markers of risk or the onset of dementia years in advance of the expression of symptoms or of a clinical diagnosis can provide sensitive outcome measures for a study of early dementia pathology^17–24^. Amongst the best characterised plasma proteomic markers for AD are lowered beta amyloid-(1-42) (Aβ_42_), its ratio with beta amyloid-1-40 (Aβ_42_:Aβ_40_) and raised phosphorylated tau (pTau), all of which accompany the accumulation of pathological proteins in the brain and clinical progression of AD ^17,25–27^ Concentrations of neurofilament light (NFL) (a non-specific marker of neuronal injury) and glial fibrillary acidic protein (GFAP) (which is associated with astrocyte activation) also are found to increase in plasma in early AD ^28–30^. Studies have identified changes in these plasma proteins prior to dementia diagnoses and brain amyloid positivity by PET^17,23,25,30^. Decreases in plasma Aβ_42_:Aβ_40_ may precede the diagnosis of AD by up to 20 years, total Tau by 15 years, with increases in NfL and GFAP detected closer to diagnosis^24^. These biomarkers provide the opportunity to assess the effect of mild to moderate SARS-CoV-2 in healthy populations on Alzheimer’s prodromal pathology and risk.

Raised levels of these dementia-related proteins have been reported in patients in the context of acute COVID-19^19,31,32^. However, interpretation of these studies is confounded by the potential for “reverse causation”, as many risk factors for severe COVID-19 are also risk factors for dementia. Complications of severe infections also can independently increase biomarker levels. To robustly test for relationships between SARS-CoV-2 infection and brain pathology for AD, longitudinal study designs that define changes from before to after infection are needed. Symptomatically mild-to-moderate infections are an ideal study target as they are less confounded by other factors associated with severe illness and prolonged intensive care. Furthermore, given that dementias also are associated with genetic background, aging, and health status, analyses need to be able to take these factors into account for elucidation of the independent effects of infection.

We were able to apply a design incorporating these principles to address the question of whether SARS-CoV-2 infection initiates or potentiates brain pathology associated with prodromal AD. We took advantage of the longitudinal blood sampling and clinical data acquisition in UK Biobank in combination with that study’s surveillance for SARS-CoV-2 exposure serology during the early stage of the UK COVID-19 pandemic (2020-21). Here we report results from over 1200 UK Biobank participants tested longitudinally for a range of plasma proteomic biomarkers. Our primary outcome measures were plasma levels of Aβ_42_ Aβ_40_, the Aβ_42_:Aβ_40_ ratio, pTau-181, NfL and GFAP (Quanterix Simoa assays). We were able to test for changes in these biomarkers with infection and compare these changes to those found in a demographically matched group who were sampled prospectively over the same interval but who were not infected with SARS-CoV-2. This longitudinal, prospective, matched case-control design permitted the modelling of changes in AD biomarkers with control for a variety of potential confounders including aging and baseline differences in genetic backgrounds, sex and health status between cases and controls. The extensive genetic, imaging and health data of the UK Biobank dataset allowed detailed assessment of confounds and relationships. In a subset of participants, we also were able to assess plasma levels of a selection of over 1400 proteins obtained from a high-throughput Olink Explore general proteomic platform, which provided the opportunity to assess the impact of SARS-CoV-2 infection on inflammatory proteins and proteins associated with risks of other chronic diseases ^20,33,34^.

## Results

### Study Population

We studied plasma proteomic biomarkers of AD and dementia risk, as well as proteins from a general proteomic panel, brain images and other health data that were made available from 1252 UK Biobank participants who had been recruited for the UK Biobank COVID-19 study (626 matched case-control pairs, 331 female)^10^. All participants had taken part in an imaging assessment session before the pandemic onset (baseline, May 2014 - March 2020). Participants were 46-80 years of age at this session (Fig. 1b). Individuals from these assessments who were determined to have previously had SARS-CoV-2 infection by at least one of: home-based lateral flow SARS-CoV-2 antibody test, PCR antigen (swab) test, GP records or public health records were invited for the COVID-19 imaging study (see Methods). Symptoms related to COVID-19 in cases were mostly mild, but 20 participants were hospitalised for COVID-19 or its complications. Of 289 cases for whom vaccination status could be determined, 40 had data indicating they had been vaccinated before their first positive COVID test. Cases were matched individually to eligible controls (defined based on age [+/- 6 months], sex, ethnicity [white/non-white], and location and date of initial imaging assessment [+/- 6 months]) who had no record of confirmed or suspected SARS-CoV-2 infection at the time of the repeat assessment. The COVID imaging assessments occurred during the UK COVID-19 pandemic (February 2021 - February 2022). Intervals between the two assessment sessions ranged from 12-82 months, matched for case-control pairs (Fig. 1 d,e,f). Blood samples for plasma proteomics were obtained at both imaging assessment sessions.

**Fig. 1.**
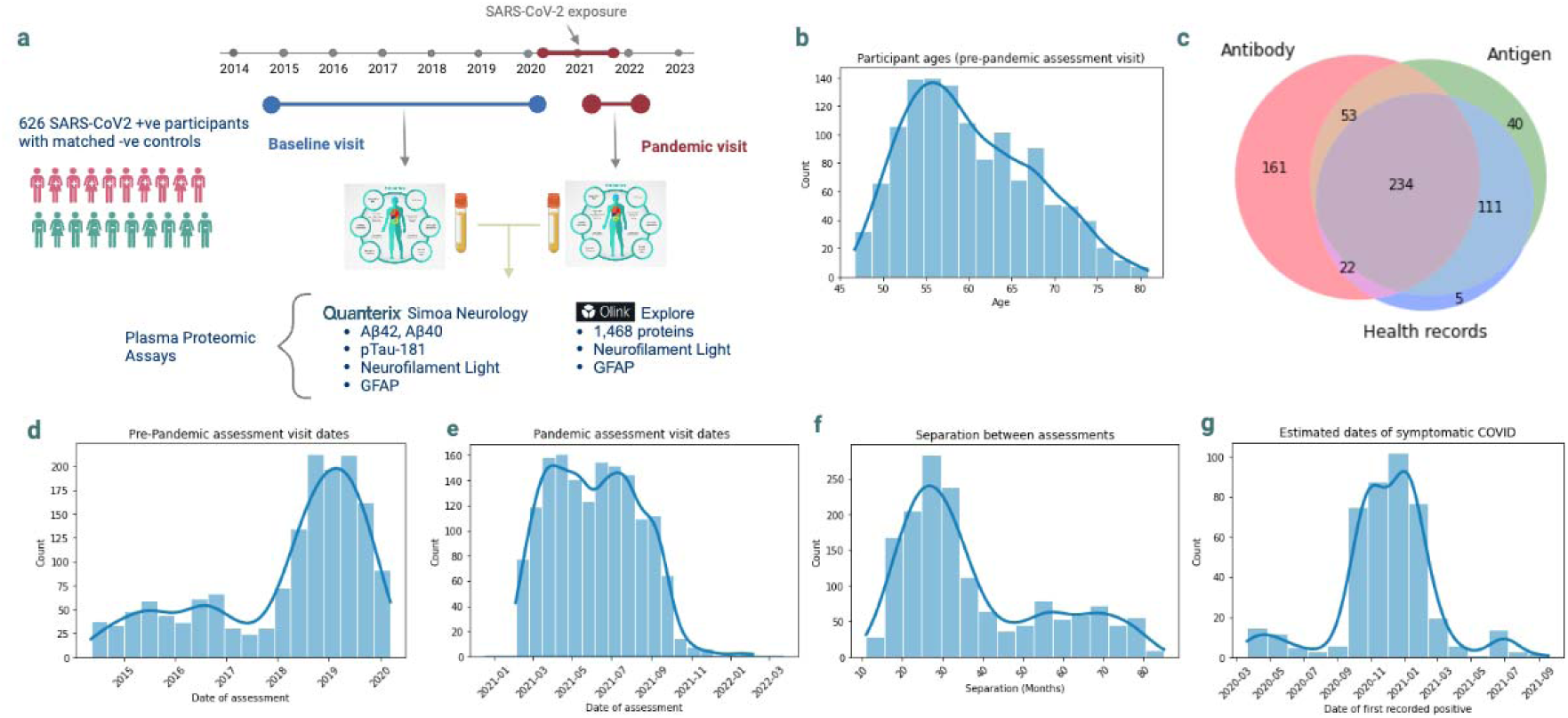
a. Experimental design. Protein concentrations were assayed from plasma samples acquired from the UK Biobank imaging assessment visits, the second of which was specifically recruited for the study of COVID-19. b. Distribution of participant ages at the pandemic assessment. c. Sources of evidence for case selection. Antibody – home-based lateral flow SARS-CoV-2 antibody test; Antigen – PCR antigen (swab) test; Health records: GP and/or hospital records d. Distribution of pre-pandemic assessment visit dates e. Distribution of pandemic assessment visit dates f. Distribution of intervals between assessments. g. Estimated dates of COVID symptoms (from participants with antigen test results).

**Fig. 2.**
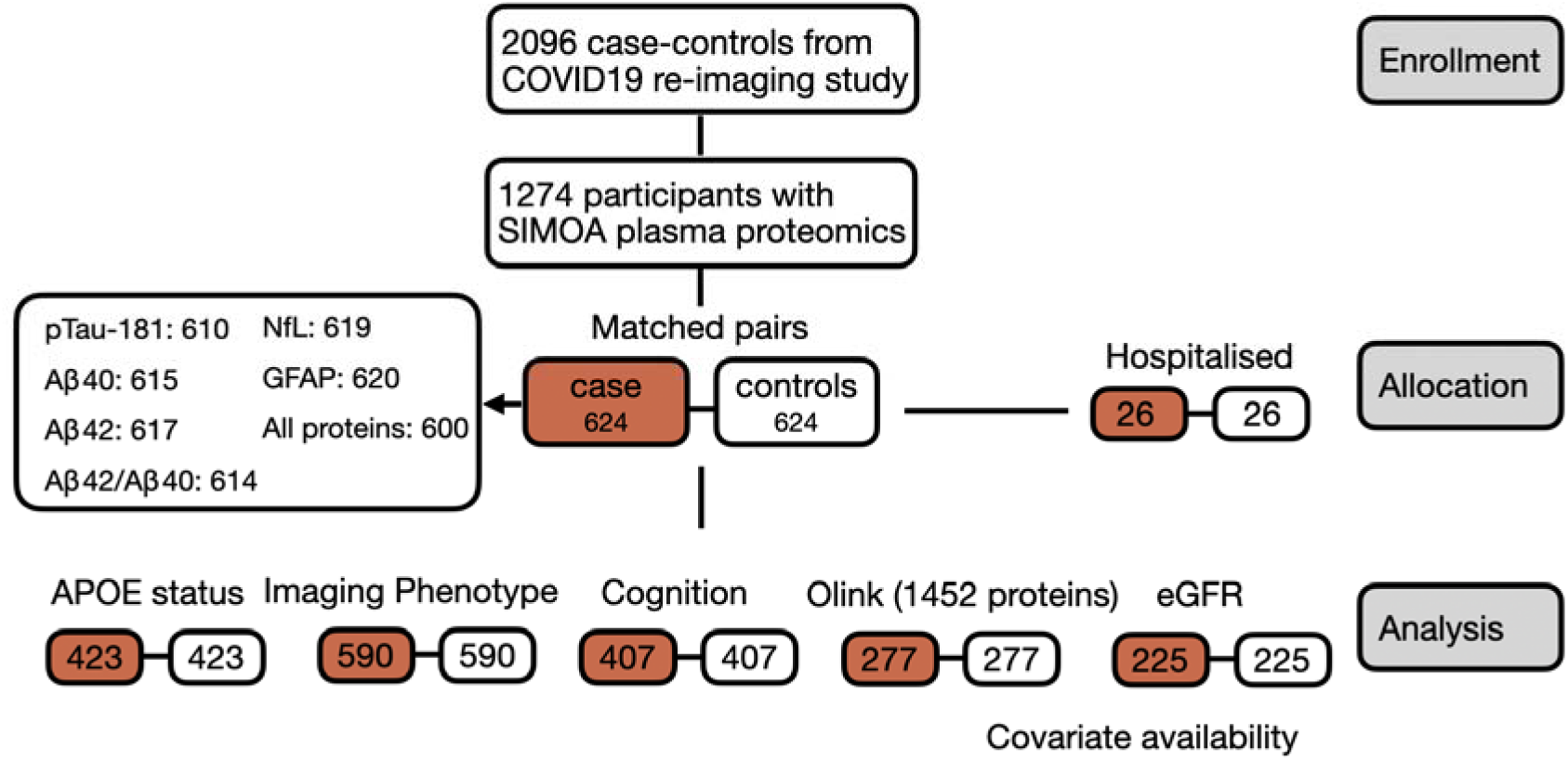
Schematic of data availability for the study. Primary analyses focus on 624 individually matched cases and controls from the COVID19 imaging repeat study with SIMOA plasma proteomic data. Full details of participant selection and control matching for the COVID study can be found at biobank.ndph.ox.ac.uk/ukb/ukb/docs/casecontrol_covidimaging.pdf.

We characterised cases and controls for potential comorbidities and other factors (Table 1). Some characteristics differed between case and control groups, reflecting lifestyle factors that could increase the likelihood of early infection: more were employed prior to the pandemic (395 vs 364); more cases (n=18) than controls (n=6) identified as “key workers” during the pandemic (p=0.023); household sizes were larger on average (p=0.007*) and cases were more active on average (p=0.002*). Cases were slightly heavier (1.4kg) than controls (p=0.028) (although hip/waist ratio and obesity rates did not differ). Other AD comorbidities did not show statistically significant differences across cases and controls. Cases had slightly higher numbers in some factors including smoking and reported prior hypertension.

**Table 1.**
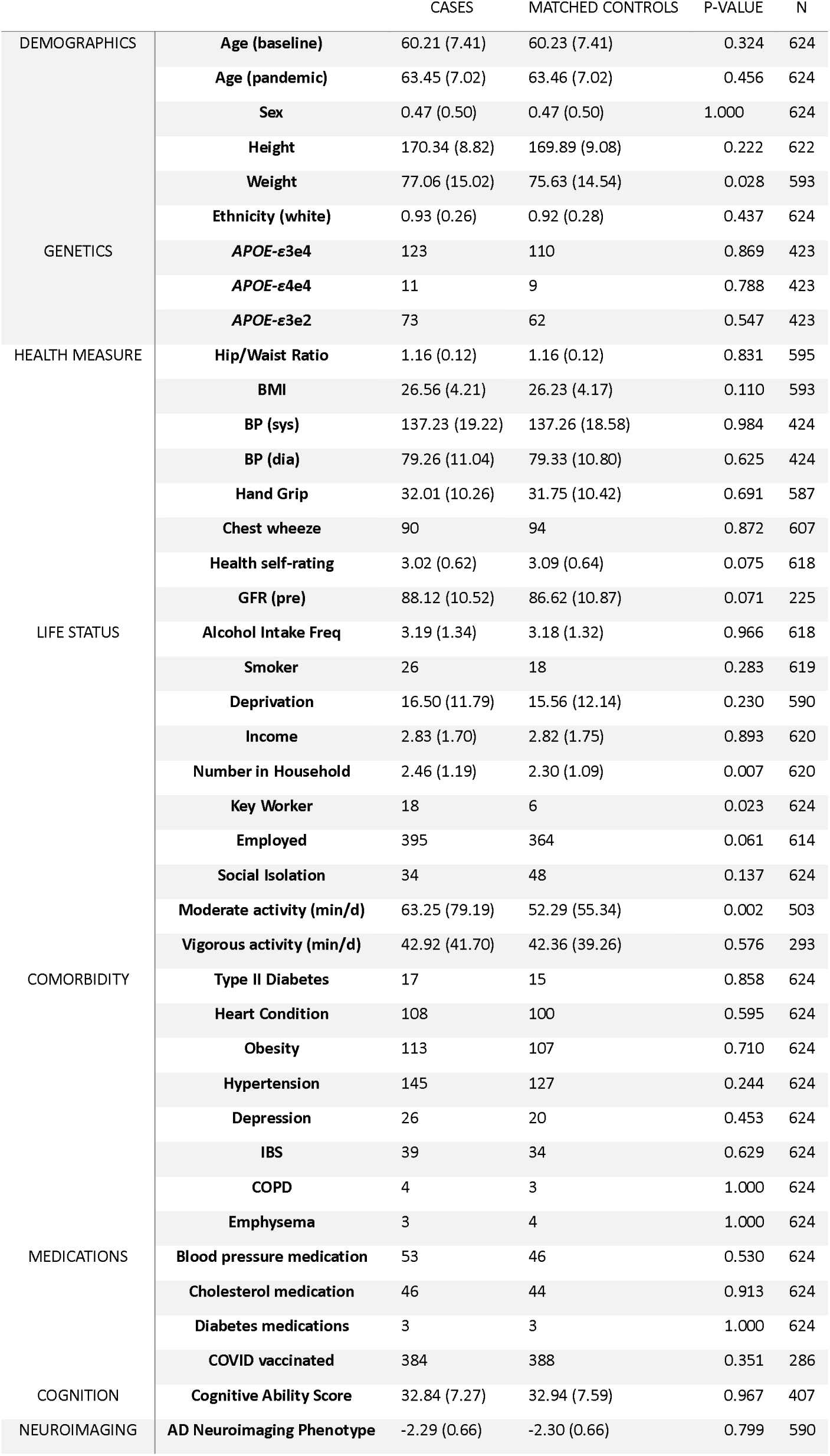
Comparisons of baseline characteristics of cases and controls. P-values reflect paired t-tests across matched case and control pairs (uncorrected). BMI: Body Mass Index, BP: blood pressure, BP medication: Prescribed anti-hypertensive medications. UK Biobank fields and derivation of measures can be found in Supplementary table 1 and Online Methods.

Single molecule array (Simoa) ultrasensitive measures of plasma amyloid-β_42_ (Aβ42), amyloid-β_40_ (Aβ42), pTau-181, glial fibrillary acidic protein (GFAP) and neurofilament light (NfL) were available on plasma collected before and after onset of the COVID pandemic for cases and for controls (see Online Methods). A complete set of longitudinal data was available for 600 cases and for 600 matched controls; the remaining cases and controls were missing data from one or more assay. Olink antibody-based proximity extension assay proteomic measurements of concentrations of 1452 proteins (including additional assays for NFL and GFAP) in the same plasma samples were available for 277 of the case-control pairs^35^.

### The plasma Aβ42:Aβ40 ratio is reduced after SARS-CoV-2 infection

To test for effects of SARS-CoV-2 infection on preclinical AD brain pathology, Twhether infection status affected longitudinal changes in concentrations of plasma Aβ40, Aβ42, pTau-181, NFL and GFAP, with the baseline assessment completed before the COVID-19 pandemic and the repeat assessment made during the pandemic, after cases had developed serological evidence for SARS-CoV-2 infection^17,36^. In addition to the plasma concentrations, we also assessed the Aβ42:Aβ40 concentration ratio^26,27^.

We fit linear models describing each participant’s pandemic-session biomarker levels in terms of their baseline, pre-pandemic levels, time interval between the two sample acquisitions, age, sex, and their individual case (serology positive, post SARS-CoV-2 infection) or control (serology negative, no evidence of prior SARS-CoV-2 infection) status (see **Error! Reference source not found.**). These models therefore modelled changes in the plasma biomarker levels between assessments, with the case-control regressor identifying differences in these changes associated specifically with SARS-CoV-2 infection. We fit further models assessing the associations of a variety of comorbidities and other factors with biomarker levels at baseline and longitudinally (Online Methods, Supplementary table 1).

Average participant plasma concentrations of Aβ42, the Aβ42:Aβ40 ratio, and pTau-181 decreased between pre-pandemic and pandemic assessments for all participants (cases and controls), while concentrations of Aβ40, NfL and GFAP increased (all p<0.0001, paired t-tests). Statistical testing of the case-control model parameters found SARS-CoV-2 infection was associated with a significantly greater reduction in the Aβ42:Aβ40 ratio (2% drop from baseline, False Discovery Rate [FDR] significant **p-value, p=0.001) (Fig. 3, Supplementary table 2). These results were maintained when an extended model including potential confounders was used (Supplementary table 3). The estimated effect of SARS-CoV-2 on the Aβ42:Aβ40 ratio was comparable to the effect of 4 years of aging (-0.5% change in Aβ42:Aβ40 ratio per year of age, estimated at baseline), and around half the average effect of heterozygosity for *APOE-ε4* (*APOE-ε4* heterozygous participants showed a 3.9% lower Aβ42:Aβ40 ratio relative to *APOE-ε3* homozygous in the baseline assessment sessions). Other comparable comorbidities of AD did not show a statistically significant associations with the Aβ42:Aβ40 ratio in this dataset. Greater reductions in the plasma Aβ42:Aβ40 ratio amongst cases were associated with greater severity of symptomatic infections: cases who were hospitalised with COVID-19 showed over twice the magnitude of reduction relative to non-hospitalised cases (5.3% vs 2.1%).

**Fig. 3.**
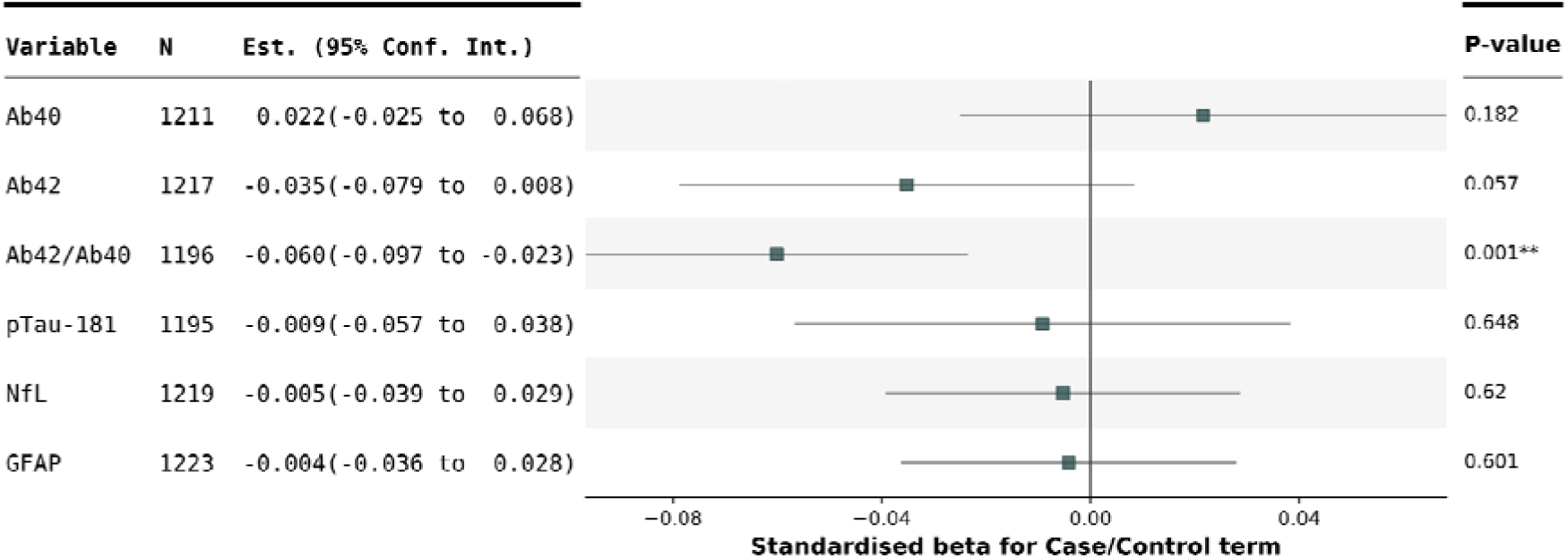
Standardised model parameters for Case/Control term in each of the AD-protein change models. Parameters represent the SARS-CoV-2 infection associated effect on change in protein levels between pre-pandemic and pandemic assessment sessions. Aβ42:Aβ40 ratios showed significant relative reductions in SARS-CoV-2 individuals. P-values correspond to one-sided tests in the directory of previously reported biomarker associations with AD. Full model fits are in Supplementary tables 2 and 3 (basic and extended models).

### Greater changes in both plasma Aβ42 and pTau-181 after SARS-CoV-2 infections were found in older participants

Participants who were older or had prior lifestyle or health conditions that are risk factors for neurodegenerative disease are likely to be more vulnerable to infection-related neurodegenerative pathology. Age is a key risk factor both for AD and for the severity of COVID-19 but the relationships between age and risk are non-linear as older people become more vulnerable to severe disease symptoms in later years^37^. To test for specific effects of SARS-CoV-2 on plasma protein level changes in older, more vulnerable study participants, we used regressors reflecting an age-related vulnerability score derived from observed associations between age and severe neurological outcomes for COVID-19 (Fig. 4c)^37^. This score has previously been used to analyse SARS-CoV-2 changes in MRI measures^10^. We assessed models that included this score and its interaction with case-control status to assess whether age-related-vulnerability predicted changes in AD-proteins between assessments with or without dependence on SARS-CoV-2 infection status (Online Methods).

**Fig. 4.**
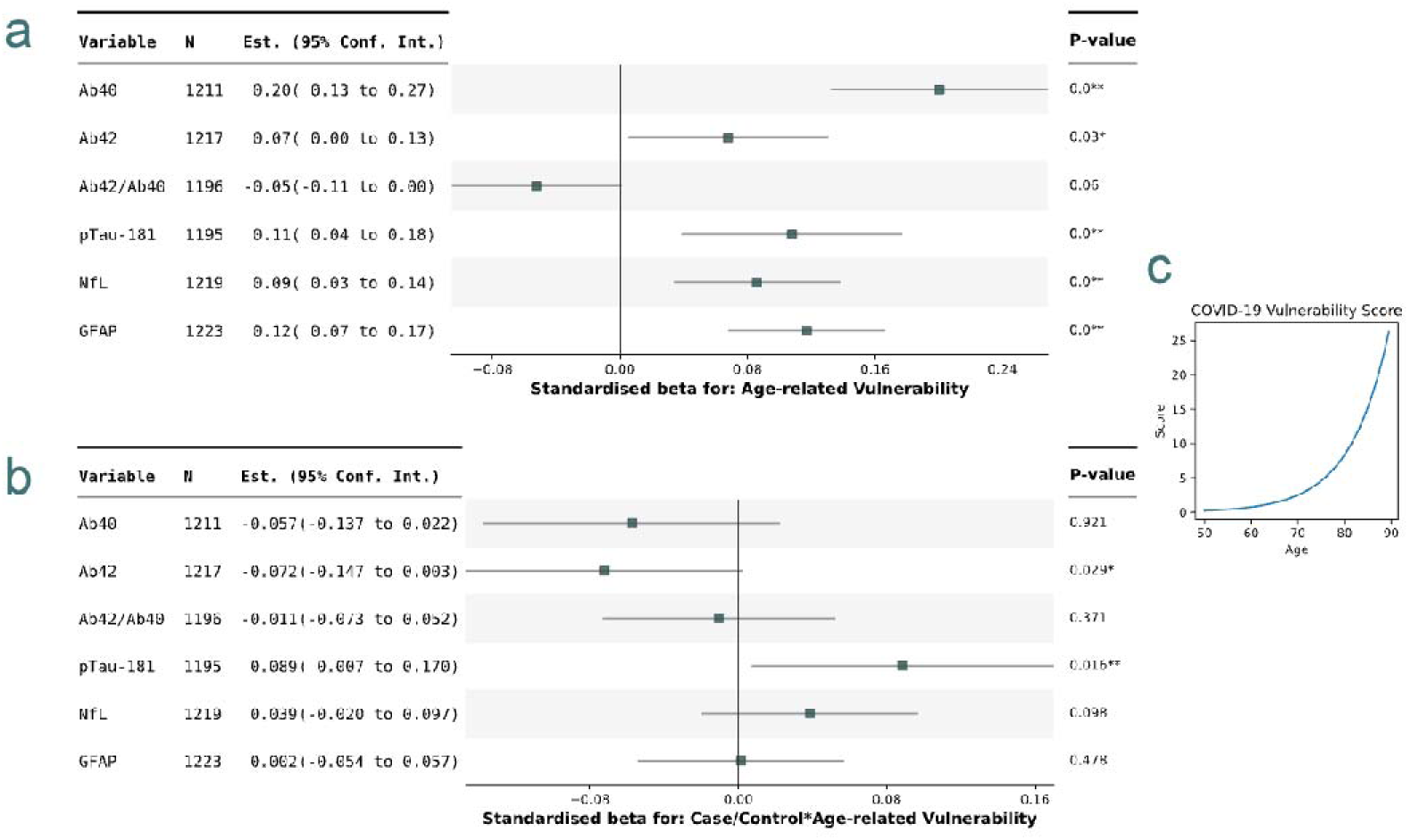
Standardised model parameters for models including age-dependent vulnerability terms in each of the AD-protein change models. a. parameters for age-dependent vulnerability term. b. parameters for age-dependent vulnerability term interaction with case-control status. c. Age-dependent vulnerability term weightings P-values correspond to one-sided tests corresponding to previously reported associations with AD. Full model fits are in Supplementary tables 4 and 5 (basic and extended models).

Across both cases and controls, a higher age-related-vulnerability score was associated with greater decreases in Aβ42:Aβ40 ratio and greater increases in pTau, NfL and GFAP concentrations between the pre-pandemic and pandemic assessment sessions (Fig. 4a, Supplementary table 4). Assessing the interaction of age-related-vulnerability score and case-control status found age-related-vulnerability score dependent increases in pTau-181 (p=0.016**) and decreases in Aβ42 (p=0.029*) in those exposed to SARS-CoV-2 relative to controls (Fig. 4b, Supplementary table 4). These effects were strengthened when an extended set of confounds were modelled (Supplementary table 5). Age-resolved plots indicate that SARS-CoV-2 cases begin to show greater changes in AD proteins from around 70 years of age (Fig. 5). For an average 75-year-old participant, this model estimates an additional 4% increase in pTau-181, a 3% decrease in Aβ40 in SARS-CoV-2 positive participants, alongside a 5.5% change in Aβ42:Aβ40 ratio (Online Methods).

**Fig. 5.**
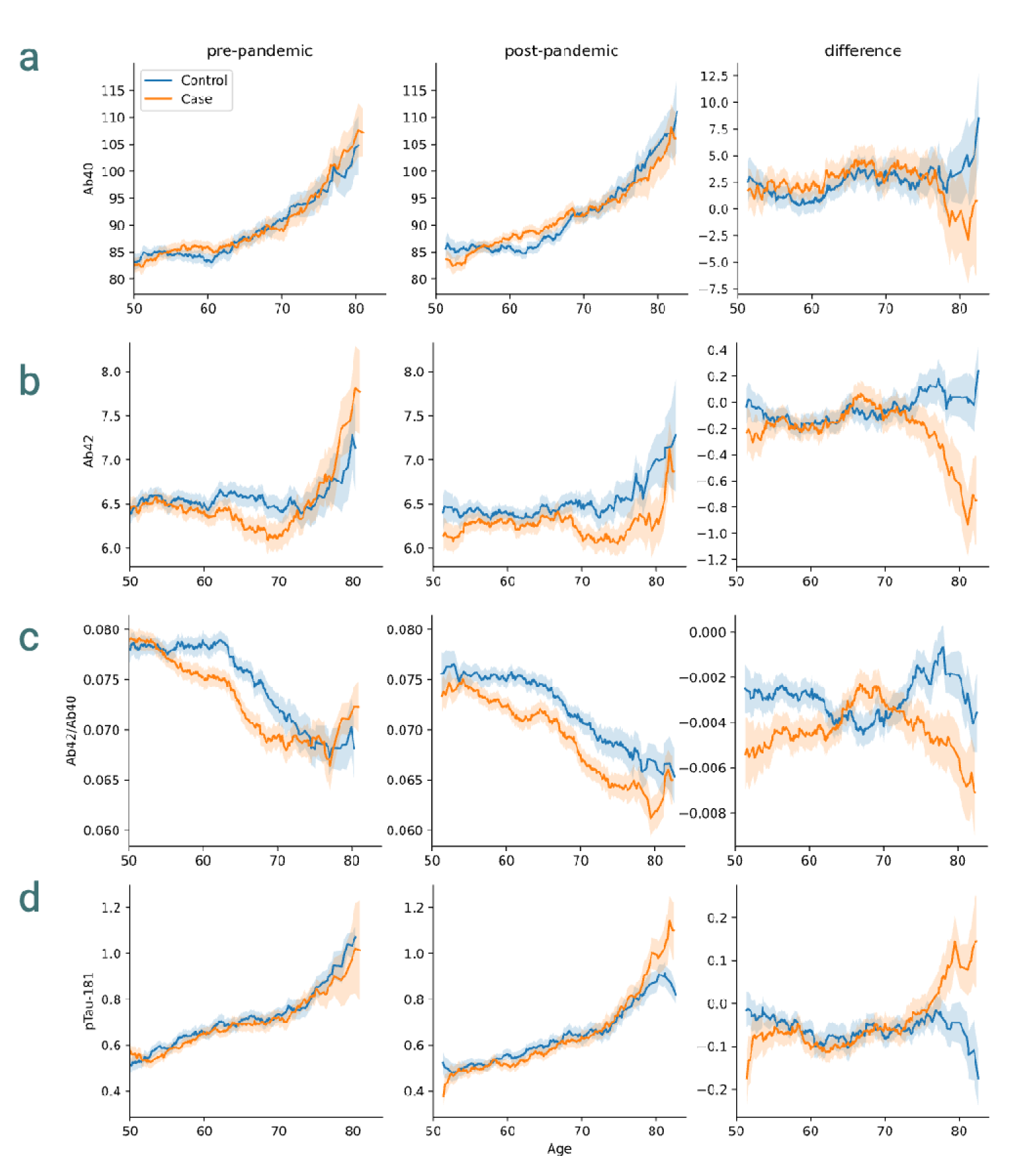
Time plots SARS-CoV-2 for plasma protein levels a. Aβ40 b. Aβ42 c. Aβ42:Aβ40 d. pTau-181. Plots show 7-year rolling mean with 90% CIs on the mean.

### AD risk factors associated with accelerated changes in plasma protein biomarker levels in cases and controls

We assessed similar models to examine the influence of a variety of further comorbidities for AD and other factors on the protein biomarkers (Supplementary table 1). Within assessment sessions, AD proteins showed rich set of associations with comorbidities and other factors (Fig. 6a, Supplementary table 6). A lower Aβ42:Aβ40 ratio was associated individuals with an *APOE-ε*4 variant (p=0.001**) and current smokers (p=0.015*). While age was matched across cases and controls, rates of these variates varied slightly across cases and controls (Table 1) and contributed to a difference in pre-pandemic Aβ42:Aβ40 ratios between cases and controls (0.0747 vs 0.0765, p=0.0145) (Supplementary table 7). These differences were attenuated by regressing *APOE* and Smoking status. While differences in baseline would not directly affect the longitudinal analysis of SARS-CoV-2 associated differences, including these factors in our primary models indicated that these effects were not confounding observed SARS-CoV-2 differences (Supplementary table 8).

**Fig. 6.**
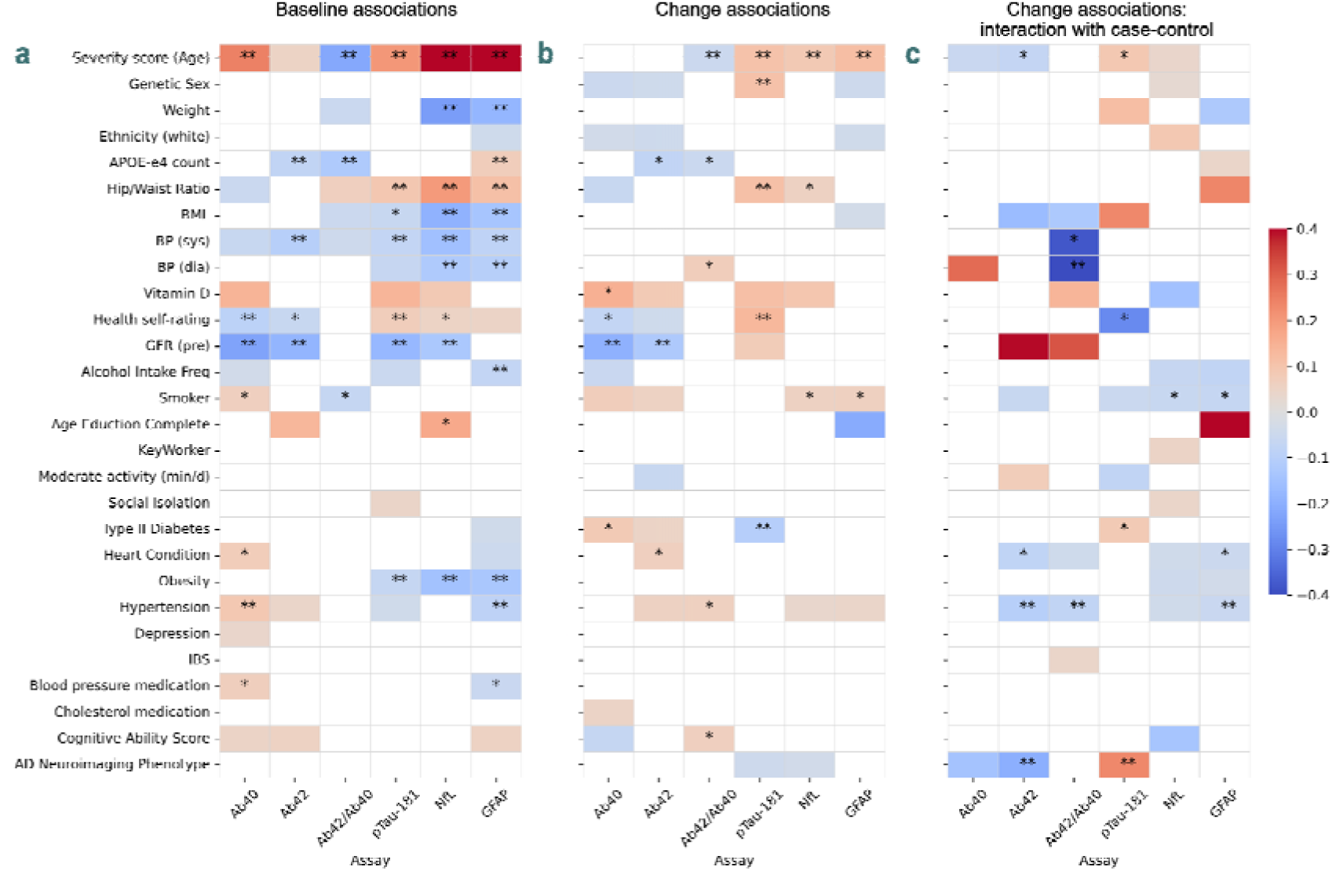
Associations between AD protein levels and comorbidities and other factors, as determined from regression models across all cases and controls. Shading shows the magnitudes of standardised betas, where model parameter fits in models controlling for age, sex and time interval between assessments. a. Associations within baseline assessment data only. Red colours represent positive associations. b. Associations of factors with the change in AD protein levels across assessments. Red colours represent a positive association of factors with levels of protein biomarker being higher in the post-pandemic assessment session. c. Interaction of association from (b) with case/control status (case=+1). Red colours represent a greater positive association in cases. (b). *: p<0.05 **: p<0.05 FDR corrected. Maps masked at abs(p)=0.20. BMI: body mass index, BP: blood pressure, GFR: estimated glomerular filtration rate, IBS: irritable bowel syndrome, pTau-181: phosphorylated Tau 181, NfL: neurofilament light.

Certain prior factors were associated with the extent of longitudinal changes in the AD proteins ( Fig. 6b, Supplementary table 9, Online Methods). Independent of case-control status, male sex, higher hip/waist ratio, and poorer health self-rating were associated with greater increases in pTau-181 across assessments (p<0.05**). Type II Diabetes appeared to be associated with reduced changes. Analysis with a model accounting for these potential confounds indicated that these differences could not account for the differences in protein levels associated with SARS-CoV-2 infection seen above (Supplementary table 10).

A low Glomerular Filtration Rate (GFR) indicates poor kidney function and is known to influence plasma protein levels^39^ and alter Alzheimer’s disease biomarker protein levels without increasing disease risk^38^. Here, lower estimated GFR (available for n=225 cases) was associated with significantly increased plasma concentrations of protein levels at baseline (Fig. 6a). Estimated GFR did not show a significant difference between cases and controls (Table 1). Higher GFR was associated with greater reductions in Aβ42 and Aβ40 protein concentrations between assessments, but not with changes in Aβ42:Aβ40 ratio (Fig. 6b). When GFR was included as a confounding covariate in models of AD biomarker change, it had negligible impact on effect sizes for SARS-CoV-2-associated differences in Aβ42:Aβ40 (Supplementary table 11).

### Prior hypertension, brain structural features associated with AD and plasma inflammatory protein levels were associated with greater SARS-CoV-2 infection-related changes in plasma AD biomarkers

Several factors showed specific associations with changes in AD-biomarker levels between assessment sessions in SARS-CoV-2 positive participants (Fig. 6c, Supplementary table 12). Hypertension was associated with greater reductions in amyloid markers of AD pathology in cases positive for SARS-CoV-2 infection (diastolic blood pressure: reduced Aβ42:Aβ40 ratio p=0.002**; hypertension: reduced Aβ42:Aβ40 ratio p=0.0018*; reduced Aβ42 p=0.004**). Hypertension has been linked to both more severe COVID-19 symptoms^39^, and the progression of AD fluid biomarkers^40^. While *APOE* genotype has been associated with COVID-19 severity previously ^44,45^ our analyses showed no clear effects of prior vaccination (p=0.44) or *APOE* status on plasma protein changes from pre- to post-pandemic in cases compared to controls. Sex also did not have a significant effect on the AD plasma proteomic changes.

A brain imaging signature was developed previously from MRI volumetric and cortical thickness measures in the Alzheimer’s Disease Neuroimaging Initiative (ADNI) database ^41^, and shown to be associated with higher rates of AD and lower memory and cognitive scores among UK Biobank participants^41^. The AD signature values were standardised for sex, age, and intracranial volume, and showed no case-control differences at baseline (p=0.779) or significant associations with AD protein levels prior to SARS-CoV-2 infection (all p>0.1). However, we found that higher values of this signature at baseline were associated with greater increases in pTau-181 (p=0.012**) and greater reductions in Aβ42 (p=0.015**) after SARS-CoV-2 infection relative to the matched controls (Fig. 6c, Supplementary table 13).

We also assessed whether baseline levels of a set of Olink inflammatory proteins associated with COVID or AD were associated with neurodegenerative biomarker changes (Supplementary table 14). None of these protein levels differed between cases and controls at the pre-pandemic session (Supplementary table 15). Higher baseline levels of Macrophage Inhibitory Factor (MIF) predicted greater reductions in Aβ protein levels across both cases and controls (Supplementary table 16). Higher baseline levels of YKL-40 (or chitinase-3 like-protein-1) were associated with greater Aβ42:Aβ40 reductions specifically in the SARS-CoV-2 group (standardised beta = -0.08, p=0.006**) (Supplementary table 17). YKL-40 is highly expressed in astrocytes during neuroinflammation and is associated with increased brain amyloid and AD symptoms^42,43^.

### Associations between changes in plasma AD biomarkers and Olink protein levels

Peripheral inflammation is a probable component of a pathway for SARS-CoV-2 induced amyloid pathology. While in mild cases of COVID-19 inflammatory protein levels have been reported to normalise in the time frame of the present study (median time since last SARS-CoV-2 positive test = 68 days)^46^, it is possible that some persistent inflammation, such as that seen in long-COVID, plays a role in the AD-related pathology we observe. Of 13 proteins assessed, TRAIL protein (TNFSF10), an apoptotic TNF cytokine was significantly reduced after SARS-CoV-2 infection (p=0.002**) (Supplementary table 18). This protein has been reported to be lower in patients with severe COVID^47^ and lower levels have been identified as a biomarker associated with AD risk in separate UK Biobank plasma samples and disease records^20^. Inflammation would be expected to be higher in participants with severe symptoms. Those participants who had been hospitalised with COVID-19 showed increased levels of Interleukin 6 (p=0.01*) and the long pentraxin PTX3 (p=0.012*) in the pandemic assessment relative to controls.

Using disease reports from the health records of 47,600 of the UK biobank participants^20^, the Olink proteins have previously been used to estimate weightings providing individual disease risk scores (ProteinScores) (online Methods). We were able to estimate 19 of these scores for the pre-pandemic and post-pandemic assessment sessions to assess whether they were associated with changes in AD-biomarkers. There were no differences in disease risk scores between cases and controls at baseline, and the incidence of all disease risks (except endometriosis) increased with age. SARS-CoV-2 was associated with increases in disease risk scores for several co-morbidities of Alzheimer’s disease: type 2 diabetes (p=0.008**), chronic obstructive pulmonary disease (COPD) (p=0.001*), ischaemic stroke (p=0.0001*) and heart disease (p=0.011*) (Supplementary table 19). These changes are consistent with recent epidemiological reports of increased incidence of some of these diseases after the pandemic ^48^. The AD risk score, weighted heavily to NfL and GFAP levels, did not show a significant increase (p=0.108).

### Associations between AD biomarker changes and changes in neuroimaging, cognitive performance, and general health measures

Previous work using the UK Biobank brain imaging data identified changes in brain structure following SARS-CoV-2 infection, linked to age-related-vulnerability^10^. We assessed whether there was evidence that these changes could be associated specifically with the AD-related changes we observe. An interaction analysis with the age-dependent vulnerability score showed evidence (p=0.027) for increases in the Alzheimer’s Disease Neuroimaging Initiative (ADNI)-derived AD structural brain phenotype score after SARS-CoV-2 infection in older participants. Increased AD brain signature scores were associated with increased pTau-181 and NfL between visits (p=0.015* and p=0.031, respectively).

Recent work has highlighted that COVID-19 is associated with long-lasting impairments in multiple cognitive domains^12^. To test for relationships between cognitive performance and SARS-CoV-2 infection in this population, we evaluated a measure of general cognitive ability derived from UK Biobank cognitive tests (see Methods)^49^. SARS-CoV-2 infection was associated with an additional 2.05% reduction in this measure over controls, equivalent to almost two years of age-related decline (1.16%/year, p=0.029). However, these changes were not strongly correlated with the AD biomarkers, which can change years prior to cognitive changes. Reductions in cognitive scores were associated with reductions in Aβ42 (p=0.037) and increases in plasma Tau-181 (p=0.011*).

Finally, participants were also asked to rate their overall health between poor and excellent. There were no significant differences at baseline between cases and controls. However, cases showed greater deterioration in their overall health ratings (2.3% p=0.007*) between the assessment visits than did the controls. While prior health self-ratings affected pTau-181 (above), changes in general health self-ratings across were not associated with AD protein levels.

## Discussion

SARS-CoV-2 infections can have a wide range of profound and long-lasting effects on human health. There have been persistent raised levels of illness and mortality since the pandemic, even in people who experienced only clinically mild cases of COVID-19^12,48,50,51^. However, evidence for the effect of COVID-19 on risk for late-life dementia has been lacking. Here, using longitudinal ultrasensitive Simoa platform plasma Aβ and pTau measures that are relatively specific and sensitive for AD and a demographically matched case-control design, we discovered plasma proteomic biomarker evidence for prodromal AD pathology after SARS-CoV-2 infection amongst UK Biobank participants. These people largely experienced mild to moderate symptoms. Despite this, SARS-CoV-2 infection was associated with significant reductions in Aβ42:Aβ40 between baseline and pandemic assessment visits. Moreover, decreases in plasma Aβ42 and increases in pTau-181 were seen in older participants and those with MRI brain signatures of AD. These protein biomarker changes have all previously been associated with increased AD risk^17,26,36,52^. While some of these changes have been associated previously with severe COVID-19 in acute settings^19,53^, to our knowledge they have not been assessed in the post-acute setting for largely mild cases. Together, our results provide evidence that SARS-CoV-2 infection initiates or accelerates brain pathology associated with AD risk and suggests increased risk of AD after infection.

Changes in plasma Aβ42:Aβ40 have been found to be the earliest detectable plasma biomarker change in preclinical stages of AD^24^ and are associated with brain amyloid accumulation even in cognitively normal participants^27^. The observed mean magnitude of the reduction in plasma Aβ42:Aβ40 after SARS-CoV-2 infection was substantial: equivalent to 4 years of aging (based on the baseline pre-pandemic data for this participant group) and 60% of the effect size of inheriting a single *APOE-ε4* allele. Effect sizes were larger in the subgroup from our study who were hospitalised for COVID-19, and for those reporting prior hypertension, suggesting that the impact on brain pathology is likely to increase with more severe infection.

We observed decreases in plasma Aβ42 and increases in pTau-181 associated with SARS-CoV-2 in participants with higher age-dependent vulnerability scores and in those with MRI brain signatures of AD prior to infection. In prior studies, concordant changes in Aβ42 and pTau-181 follow Aβ42:Aβ40 in the progression of plasma biomarkers in pre-AD ^24,25^ and have been associated with increasing brain Aβ load ^17,26,36,52,54^. These changes in more vulnerable participants were associated with brain structural changes, increased cognitive deficits, and decrements in measures of general health quality.

Increases in NfL and GFAP in CSF and plasma have been reported previously with Alzheimer’s disease, MCI, and acute COVID ^17,26,53,55,56^. Here we did not find increased levels in either following SARS-CoV-2 infection. NfL and GFAP are markers of neural injury and astroglial activation, respectively, which are more predominant in later stages of AD progression. While the plasma Aβ42:Aβ40 ratio have been found to decrease from control levels up to 18 years prior to an AD diagnosis, increases in plasma NfL and decreases in hippocampal volume were not detectable until at least 9 years later^24^. We would expect few participants to have such progressed AD pathology. The absence of changes in these proteins also aligns with observations made during and after COVID. In hospitalised COVID patients, plasma NFL and GFAP concentrations have been observed to be raised, but they returned to control levels within 3 months for the majority of patients ^57^.

There are a variety of mechanisms by which mild to moderate SARS-CoV-2 infection may increase the brain beta-amyloid load. Brain beta-amyloid production and aggregation can be potentiated by stress, infection and pathological conditions including diabetes, stroke and vascular disease ^58,59^. Epidemiological studies have suggested that a wide range of systemic infections may be associated with increased risks of AD ^60,61^. SARS-CoV-2 is not neurotrophic^62^, but infection is associated with systemic inflammatory activation^63^. With associations between peripheral inflammatory events and dementia identified in numerous studies^64^, possible neuroimmune mechanisms of AD pathogenesis are increasingly being studied ^65–67^. Pre-clinical studies of peripheral inflammatory challenges have found evidence both for potentiation of brain microgliosis and for impairment of microglial clearance of beta-amyloid and the progression of tau pathology ^7,68^ Levels of midlife systemic inflammatory markers have been associated directly with late-life relative brain atrophy comparable to that associated with the *APOE4* allele^69^, an effect size mirroring that observed here.

Peripheral inflammation may influence CNS inflammatory processes in several ways. For example, systemic infection can enhance CNS inflammation directly by trafficking of activated immune cells or indirectly with increases in systemic cytokines or inflammatory activation of endothelial cells and breakdown of the blood-brain-barrier to allow leak of chronically pro-inflammatory plasma proteins into the CNS^14,15^. Direct infection of vascular endothelial cells by SARS-CoV-2 would enhance this ^70^. Long-lasting immunological changes in patients could maintain these effects for prolonged periods even in mild-to-moderate cases^71^. One specific mechanism is increased brain interferon-induced transmembrane protein (IFITM1-3) expression triggered by viral infection. Increased incorporation of IFITM3 into the g-secretase complex potentiates its activity, increasing Aβ production ^72,73^.

The AD biomarker proteins studied here are being increasingly assessed at scale^24,74^, but their patterns of their variation in the general population are not fully characterised. Aβ and pTau are strongly linked to AD and neurodegeneration, but they can vary with specific conditions other than AD^87^. In the present dataset, concentrations of these proteins showed associations with a variety of traits including hypertension, weight, GFR and Type II Diabetes diagnosis. However, the observed SARS-CoV-2 infection-related effects remained when these factors were included in the model as potential confounders.

In addition to highlighting increased plasma biomarkers of prodromal AD with SARS-CoV-2 infection, application of the more general Olink plasma protein-based risk scores developed in an earlier UK Biobank study also provides evidence for greater predicted risks for diabetes, ischaemic stroke and heart disease, COPD, lung cancer and endometriosis in the subset of participants under the age of 73^20^. This is consistent with emerging post-COVID-19 epidemiology data ^76–79^ and proteomic studies of samples from COVID-19 patients hospitalised with acute symptoms^19^. While we found limited evidence for comorbidities directly confounding our AD biomarker observations, the raised risk of many - albeit not all – diseases with SARS-CoV-2 infection raises the possibility that SARS-CoV-2 (and other infections) may accelerate both body and brain aging ^82^. The changes in these risk-markers across diseases may be due to common (e.g., inflammation) processes initiated with SARS-CoV-2 infection, a hypothesis supported by findings of associations of Alzheimer’s disease with multiple co-morbidities^88^. It is likely that infections can have both general and specific impacts on disease risks, and will affect different organ systems^1,23^.

Patients with hepatic dysfunction have raised levels of plasma beta-amyloids and increased Aβ42/40 ratios^82,83^. However, this is opposite to the direction of change observed here. We also did not find evidence for increases in Olink protein signatures associated with hepatic dysfunction after SARS-CoV-2. Kidney function can also affect plasma protein levels. We identified associations between eGFR and protein levels, but these did not confound our primary results. Increased plasma pTau-181 is associated with combined peripheral and central nerve pathology in ALS and with spinal muscular atrophy^76^. While further research is necessary^77^ , we believe that the association of both increased plasma pTau-181 and decreased plasma beta-amyloid 42/40 ratios with SARS-CoV-2 infection most likely reflects brain pathology.

While our study was able to combine fluid biomarkers, imaging data, cognitive performance, and other covariates in a longitudinal case-control design, it has limitations. We relied on data linkage to hospital records and retrospective reports for information concerning symptoms associated with SARS-CoV-2 infections, limiting our ability to investigate detailed relationships between the clinical severity of infections and plasma biomarker changes. While we used prospectively acquired samples from before and after SARS-CoV-2 infection, the pragmatic, observational nature of our analyses precludes making strong causal inferences. While case and control groups were well matched by most measures (including cognitive scores, inflammation markers and disease risks), there were some differences variables in body mass, activity levels, and diabetes mellitus and hypertension rates. While analyses controlled for these and other known risk factors, it is possible there are additional, uncontrolled susceptibility factors that distinguished the SARS-CoV-2 infection and control groups. Asymptomatic cases may be under-represented in the sample. This does not invalidate the assessment of accelerated changes in biomarkers in the current sample relative to baseline levels, but it means that these changes may be lower in the broader population. Generalisation of our results to the wider population must be done with care: the UK Biobank sample population is not representative of the general population by many measures^83^.

Recent studies have compared various blood phosphorylated tau immunoassays, notably those measuring pTau-217 and -231, for their utility as biomarkers for AD^84^ . In head-to-head assays, all were able to identify AD and brain Aβ pathology^85^. pTau-217 has shown a greater sensitivity to later-stage disease progression, but pTau-231 and pTau-181 may show greater sensitivity to early stage amyloid accumulation, the primary disease phase of interest here ^87,88^.

In conclusion, our study, along with earlier studies suggesting increased rates of dementia diagnosis following COVID-19^89^, suggests that public health planning should take into account the possibility of increased rates of AD (and other systemic diseases) in the coming years due to the COVID-19 pandemic. Moreover, the relationships described here are unlikely to be specific for SARS-CoV-2 and may reflect more general effects of systemic viral or other infections on the aging brain. This highlights the need to prioritise pandemic preparedness as part of a strategy for reducing the future long-term burden of dementia and other chronic diseases, as well as for the direct impact of the acute infections.

## Online Methods

### Ethics

The UK Biobank is approved by the North West Multi-Centre Research Ethics Committee (MREC) to obtain and share data and samples from volunteer participants. Written informed consent was obtained from all participants (http://www.ukbiobank.ac.uk/ethics/).

### Design

This study analyses Simoa ultra-sensitive assays of dementia-related proteins taken from blood-plasma obtained from participants in the UK Biobank COVID-19 repeat imaging study)^10^. This study identified case and control participants from those who had taken part in the UK Biobank imaging enhancement^90^, which comprises a comprehensive imaging assessment at one of four dedicated sites. Over 40,000 participants had been assessed prior to the COVID-19 pandemic. Inclusion criteria and other details are described in https://biobank.ndph.ox.ac.uk/showcase/showcase/docs/casecontrol_covidimaging.pdf.

Participants were identified as being SARS-CoV-2-positive by the UK Biobank if (a) there were positive diagnostic antigen tests identified in the linkage to health-related records, (b) COVID-19 was reported in their primary care data or hospital records, or (c) two home-based lateral-flow antibody tests provided by the UK Biobank were positive (Fortress Fast COVID-19 Home test, Fortress Diagnostics and ABC-19TM Rapid Test, Abingdon Health). Both diagnostic antigen test and GP/hospital records were accompanied by data permitting estimation of date of COVID-19 infection.

In all, Simoa biomarker measurements from 626 cases and their matched controls were available. Each participant had biomarker measurements from blood samples taken longitudinally from the two imaging assessment sessions (pre- and post-pandemic). Complete pairs of these data were available from 600 cases and from the 600 their matched controls had complete data. For the 626 cases, SARS-CoV-2 infection was determined from mixed sources: 358 had a general practitioner’s diagnosis of COVID-19 , 438 had record of a positive diagnostic antigen test prior to the assessment, and 467 returned positive on home-based antibody lateral flow test kits (Fig 1b). 24 participants had SARS-CoV-2 infection determined during a period of hospitalisation, for which 20 had COVID-19 listed as a primary cause. Two participants were excluded for having a prior dementia diagnosis (vascular dementia), identified from hospital inpatient records.

Control participants were selected as those who had negative antibody test results, and / or no other record of confirmed or suspected COVID-19 from available data. Control participants were matched to positive cases based on five characteristics: sex, ethnicity (non-white/white), date of birth (+/- 6 months), the location of first imaging assessment, and the date of this assessment (+/- 6 months). Further details of case and control identification are provided in the above link and ^10^.

### Proteomic assay acquisition and pre-processing

Aβ40, Aβ42, NfL and GFAP concentrations were measured using the Simoa Human Neurology 4-Plex E assay (Quanterix). The Simoa P-tau181 Advantage Kit was used to measure P-tau181 concentration. All measurements were made on an HD-X instrument (Quanterix) with one round of experiments and a single batch of reagents. Technicians were blinded to participant case-control status and from which assessment visit a sample was taken from. Measurements were all above limit of detection and coefficients of variation within analytes were below 10%.

Anonymised data from 1256 participants were integrated with phenotype data via a UK Biobank key. Four participants were excluded for data being only available for one of the two imaging sessions. P-tau181, NfL, and GFAP markers showed positively skewed distributions and were log-normalised. For each biomarker, outliers were identified as measurements more than 8 times the Median Absolute Deviation (MAD) and excluded from the analysis. PCR plate IDs were also regressed out from the data. There was no significant imbalance of outliers or plate IDs across cases and controls. Cases and controls were matched according to UK Biobank field 4100 with 609 case-control pairs identified. 1220 participants and 583 matched case-control pairs had complete proteomic data. All analyses were performed on matched data, except (as noted) when key covariates of interest were missing from a set of participants. The Simoa Neurology assays showed patterns of covariance across participants closely matching previous reports (Fig 5) ^91^.

### Olink Proteomics

We used Olink Explore proteomic data from the Pharma Proteomics Project (PPP) which assayed samples from 54,219 UK Biobank participants across the different assessment sessions ^35^. Available data from the COVID-19 imaging assessment sessions for our case/control matched pairs comprised 436 matched participant pairs for which 1474 proteins from Cardiometabolic, Inflammation, Neurology and Oncology Olink panels were assayed. The Olink 3072 Explore platform extension data were not available for this COVID-19 case-control matched assessment visits. Available Olink proteins included NFL and GFAP which were also profiled in the Simoa assays. These markers were positively correlated to their Simoa counterparts (r=0.70 and 0.53 respectively).

The Olink panel included Cystatin-C, which permitted estimation of the Glomerular Filtration Rate (eGFR) in the imaging assessment sessions using the KD-EPI Cystatin C Equation^92^. We first assessed how well protein levels measured by OLINK related to serum Cystatin-C measurements made separately and included in the UK Biobank core blood biochemistry panel data. The latter were derived from the Siemens Advia 1800 platform protein and available only for the initial UK Biobank assessment session. OLINK Cystatin-C measures in our study baseline plasma samples and those from earlier plasma samples acquired on the same participants showed a correlation of 0.84, which was used for appropriate rescaling of the OLINK data to enable eGFR estimation from the OLINK data.

We assessed 13 inflammatory proteins identified from a literature search of associations of inflammatory proteins in blood or CSF with COVID and/or Alzheimer’s disease (Supplementary table 14). The inflammatory proteins and risk scores were analysed for baseline case-control differences and associations with AD proteins change using the statistical models outlined below.

We also investigated whether recently defined multivariate plasma protein concentration changes associated with disease risk (ProteinScores) derived from Olink Explore plasma protein biomarkers in 47,600 UK biobank participants^20^ provided evidence for increased systemic chronic disease risks following SARS-CoV-2 infection. These scores were derived using penalised Cox elastic net regression which linked protein levels some samples in the initial UK Biobank assessment session to over 16 years of primary and secondary care NHS records disclosing 21 incident outcomes. Being penalised regression, each score comprises weights from 10-20 Olink proteins, selection of which was validated using hold-out data from the UK Biobank. As these risk scores were derived and validated on participants below the age of 73 years, we limited our analysis to this subset of our case-control cohort (i.e., the 353 cases and 353 matched controls under the age of 73).

We generated these risk scores for protein profile data from the COVID-19 imaging assessment sessions. The data were rank-base inverse normal transformed and rescaled to match the processing applied in the generation of the ProteinScores. Disease weight scores were downloaded from the publication website and applied to the data^20^.

### Phenotypic data processing

Primary data for the study comes from the two imaging assessment sessions. Genetic and phenotypic data, including imaging and health records were available for all participants who took part in the COVID-19 reimaging project. Symptomology analyses used data from the UK Biobank COVID-19 Serology Study^93^. Data were parsed and cleaned using the FMRIB UK Biobank Normalisation, Parsing and Cleaning Kit, and integrated with other data modalities into a combined Python pandas data frame.

### Genetics

*APOE* variant status was extracted from UK Biobank subject genotyping using PLINK-2.0^94^. We extracted variants at SNPs *rs429358* and *rs7412* to identify participants’ *APOE* status. Data was available for 1027 participants, with five genotypes: ε3ε3 (n=630), ε3ε4 (235), ε2ε3 (135), ε4ε4 (20), and ε2ε2(7). We used ε3ε3 genotype as reference when one was required. Additional AD and COVID-19 -associated single nucleotide polymorphisms in the UK Biobank dataset were sequenced directly or imputed.

### Imaging data and neuroimaging AD phenotype score

To avoid the multiple testing challenge of large numbers of neuroimaging measures and provide a measure recognised to be relevant to AD, we estimated a structural brain data AD phenotype score for each participant at each assessment session using structural brain imaging-derived measures made available through the UK Biobank Showcase^95,96^. The AD phenotype score, developed on the Alzheimer’s Disease Neuroimaging Initiative (ADNI) dataset, reflects the prediction of a Bayesian machine learning neural that a participant has AD from 155 FreeSurfer cortical volume and thickness measures^41^. Monte Carlo dropout was used to approximate Bayesian inference within a two-layer neural network. This phenotype model was trained on 736 individuals (331 AD) from the ADNI database, and validated on 5209 participants from the National Alzheimer’s Coordinating Centre and 37,104 participants in the UK Biobank ^41^, where predicted at-risk participants showed cognitive profiles representative of AD.

Neuroimaging measures contributing to the AD phenotype were processed using the same steps as used for the generation of the model. Each cortical volume and thickness measures was cleaned by removing outliers more extreme than eight times the median absolute deviation from the median (across both pre-pandemic and pandemic assessment visits). The measures were then de-confounded for age, estimated total intracranial volume, and sex, and rescaled using normalisation statistics from the ADNI training set. Software to generate these signatures was downloaded from https://github.com/tjiagoM/adni_phenotypes. PyTorch was used to apply the trained model (https://wandb.ai/tjiagom/adni_phenotypes/runs/2cxy59fk) to UK Biobank data.

### Cognitive Scores

The UK Biobank includes a series of standard cognitive tests, but they can be individually unreliable, and do not together correspond to established tests for general or dementia-related cognitive scoring. A procedure to estimate a measure of general cognitive ability from the UK Biobank data has previously been developed using an external validation dataset ^49^. We used the resulting score weights to calculate an overall score for cognitive ability for participants in each assessment visit.

### Statistical modelling

We assessed longitudinal change in proteomic biomarkers and the effect of SARS-CoV-2 on this change using linear models which modelled protein levels in the pandemic assessment samples in terms of the pre-pandemic protein concentrations, the time between assessment visits, designation as a case or control and potential confounders including age and baseline differences genetic backgrounds, sex, health status or social risk factors for COVID between cases and controls^10^. We approached this is a staged fashion that allowed the impact of confounder covariates (some which were available only in a proportion of subjects) to be understood. Our basic model included covariates for Sex, Age, and the interval between assessments:

Protein_post = Protein_pre + Interval_between_assessments +

Interval_between_assessments^2 + Case/control + Genetic_sex + Age_post

The extended models included further variables known or expected to change over the assessment period and with COVID-19 (Table 4). Some variables presented considerable missing data, such that the extended models had reduced power.

As the impact of SARS-CoV-2 infection is expected to vary according to certain characteristics and vulnerabilities, we fit further models including interaction terms modelling the interaction of SARS-CoV-2 status with characteristics including *APOE* status, hospitalisation, and pre-pandemic neuroimaging AD score, to determine if these factors modified the effect of SARS-CoV-2 on protein levels. Age is an important vulnerability factor. Past modelling studies of brain structural changes associated with COVID-19 have used a score of age-dependent vulnerability as an interaction term to account for the greater vulnerability of the brain with age^10^. An exponential age-dependent vulnerability score term (10^Age_×_0.0524_−_3.27^) was derived from the modelling of the observed relationship between age and COVID-19 neurological symptom severity^37^. All these interaction models included both the original case-control variable and the original vulnerability terms in addition to the interaction term, along with additional covariates used in the basic model.

In addition to protein levels, we modelled several additional outcome measures: disease risk scores, a general cognitive ability score, a neuroimaging AD score, and reported level of general health. These outcomes were modelled in the same manner as the protein levels above. To assess whether changes in these outcomes were associated with changes in protein scores, additional models were assessed which included as predictors the pre-pandemic levels of proteins and their change across assessment visits. Linear models were also used to assess relationships between protein levels and UK Biobank variables at baseline. Age, sex, and interval between assessments were used as confound variables in these models as above.

The models were fit using Ordinary Least Squares and tested model parameters using t-tests. As we had strong hypotheses regarding direction of SARS-CoV-2-induced change in AD protein levels based on prior literature, we used one-sided tests. Benjamini/Hochberg False Discovery Rate with alpha=0.05 was used to control for statistical testing of multiple hypothesis across proteins and/or covariates.

## Data and code availability

All data is available upon application from the UK Biobank.

Code for analyses and figures will be shared from the UKDRI GitHub page.

## Data Availability

All data is available upon application from the UK Biobank.

https://www.ukbiobank.ac.uk/

## Acknowledgements

This research has been conducted using the UK Biobank Resource under Application Number 76059. We are grateful to the participants, researchers, and all UK Biobank staff for their contributions to enabling this research. We particularly acknowledge the efforts of the staff and the commitment of UK Biobank participants to allowing the acquisition of these data during the COVID-19 pandemic. We also want to acknowledge support for the conception, development, funding and interpretation of results of the study from Dr. Sally Johns of Biogen. We thank the anonymous reviewers for their valuable suggestions. Figures were created with BioRender.com.

EPD is supported jointly by funding to PMM from the UK Dementia Research Institute and from the NIHR Imperial Biomedical Research Centre, in which he is a member of the Multiple Long-Term Conditions theme. HZ is a Wallenberg Scholar supported by grants from the Swedish Research Council (#2023-00356; #2022-01018 and #2019-02397), the European Union’s Horizon Europe research and innovation programme under grant agreement No 101053962, Swedish State Support for Clinical Research (#ALFGBG-71320), the Alzheimer Drug Discovery Foundation (ADDF), USA (#201809-2016862), the AD Strategic Fund and the Alzheimer’s Association (#ADSF-21-831376-C, #ADSF-21-831381- C, and #ADSF-21-831377-C), the Bluefield Project, Cure Alzheimer’s Fund, the Olav Thon Foundation, the Erling-Persson Family Foundation, Stiftelsen för Gamla Tjänarinnor, Hjärnfonden, Sweden (#FO2022-0270), the European Union’s Horizon 2020 research and innovation programme under the Marie Skłodowska-Curie grant agreement No 860197 (MIRIADE), the European Union Joint Programme – Neurodegenerative Disease Research (JPND2021-00694), the National Institute for Health and Care Research University College London Hospitals Biomedical Research Centre, and the UK Dementia Research Institute at UCL (UKDRI-1003). PMM acknowledges generous personal support from the Edmond J Safra Foundation and Lily Safra and an NIHR Senior Investigator Award. His work additionally is supported by the UK Dementia Research Institute, which receives its funding from UK DRI Ltd., funded by the UK Medical Research Council, Alzheimer’s Society and Alzheimer’s Research UK and, for this research specifically, Biogen.

## Supplementary Information

Tables not shown can be found in the accompanying Excel spreadsheet.

**Supplementary table 1.** UK Biobank Covariates used in analyses. BMI: Body Mass Index; AD: Alzheimer’s Disease, PRS: polygenic risk score. COPD - Chronic obstructive pulmonary disease

| CLASS | COVARIATE | UKB DATA-FIELD ID(S) | CASE/CONTROL COMPARISON | BASIC MODEL | AD RISK FACTORS MODELS |
| --- | --- | --- | --- | --- | --- |
| **DEMOGRAPHICS** | Age | 34,52,53 | X | X | X |
|  | Sex | 31 | X | X | X |
|  | Height (cm) | 50 | X |  |  |
|  | Weight (kg) | 21002 | X |  | X |
|  | Ethnicity (White) | 21000 | X |  | X |
| **ASSESSMENT INFORMATION** | Date of assessment/interval | 53 | X | X | X |
| **GENETICS** | APOE status | Genotyping | X |  | X |
| **HEALTH MEASURES** | Body Mass Index | 21001 | X |  | X |
|  | Hip/Waist Ratio | 48,49 | X |  | X |
|  | Blood Pressure (Diastolic, Systolic) | 4079,4080 | X |  | X |
|  | Hand grip strength | 46,47 | X |  |  |
|  | Chest Wheeze | 2316 | X |  |  |
|  | Est. Glomerular Filtration Rate | OLINK: P_CST3 | X |  | X |
|  | General Health | 2178 | X |  | X |
| **LIFE STATUS** | Alcohol Intake Freq | 1558 | X |  | X |
|  | Smoking status | 1239 | X |  | X |
|  | Years of education | 845 | X |  | X |
|  | Deprivation | 26410 | X |  |  |
|  | Income | 738 | X |  |  |
|  | Household Size | 709 | X |  | X |
|  | Employment status | 6142 | X |  |  |
|  | Key Worker Status | 28063 | X |  |  |
|  | Activity levels (min/day) | 894,914 | X |  | X |
|  | Isolation (social visits) | 1031,2110 | X |  | X |
| **COMORBIDITIES** | Type II Diabetes | 26206 | X |  | X |
|  | Heart Condition | 20002 | X |  | X |
|  | Obesity | 21001 | X |  | X |
|  | Hypertension | 20002 | X |  | X |
|  | Depression | 20002 | X |  | X |
|  | Irritable Bowel Syndrome | 20002 | X |  | X |
|  | COPD | 20002 | X |  | X |
|  | Emphysema | 20002 | X |  | X |
|  | Renal conditions | 20002 | X |  | X |
| **MEDICATION** | Blood pressure medication | 6153 | X |  |  |
|  | Cholesterol medication | 6153 | X |  |  |
|  | Diabetes medication | 6153 | X |  |  |
|  | COVID vaccination | Health Records | X |  |  |
| **COGNITION** | General Cognitive Ability score | Cognitive test fields | X |  | X |
| **NEUROIMAGING** | AD imaging phenotype | FreeSurfer IDPs | X |  | X |

Supplementary table 2 Parameter fits for linear model predicting proteomic levels in pandemic assessment session. Table shows model fits and p-values associated with case / control status (case=+1), age, sex (male=+1), the time interval between sessions (days), and pre-pandemic protein level. Beta estimates have been standardised. NfL – neurofilament light; GFAP (**) indicates significant, FDR corrected by column, alpha=0.05.

Supplementary table 3 Parameter fits for linear model predicting proteomic levels in pandemic assessment session with an extended set of confound variables. Beta estimates have been standardised. NfL – neurofilament light; GFAP (**) indicates significant, FDR corrected by column, alpha=0.05.

Supplementary table 4 Parameter fits for linear model predicting proteomic levels in pandemic assessment session with additional terms modelling age-related vulnerability and its interaction with case/control status. Beta estimates have been standardised. NfL – neurofilament light; GFAP (**) indicates significant, FDR corrected by column, alpha=0.05.

Supplementary table 5 Parameter fits for linear model predicting proteomic levels in pandemic assessment session with additional terms modelling age-related vulnerability and its interaction with case/control status, and an extended set of confound variables. Beta estimates have been standardised. NfL – neurofilament light; GFAP (**) indicates significant, FDR corrected by column, alpha=0.05.

Supplementary table 6 Associations of protein biomarkers with UK Biobank variables at pre-pandemic assessment visit. Associations were determined from linear models including age and sex covariates. (*) p<0.05 uncorrected (**) p<0.05 FDR correction (alpha = 0.05).

Supplementary table 7 Baseline protein level statistics for cases and controls. P-values reflect paired t-tests.

Supplementary table 8 Parameter fits for linear model predicting proteomic levels in pandemic assessment session with additional terms modelling covariates APOE variant status, smoking status, Hip/Waist ratio and Diabetes. Beta estimates have been standardised. NfL – neurofilament light; GFAP (**) indicates significant, FDR corrected by column, alpha=0.05.

Supplementary table 9 Associations of UK Biobank variables (measured at pre-pandemic assessment) with the change in protein biomarkers across assessments (post-pandemic – pre-pandemic). Associations were determined from linear models of protein level change including terms for age, sex, and interval between assessments. (*) p<0.05 uncorrected (**) p<0.05 FDR correction (alpha = 0.05).

Supplementary table 10 Parameter fits for linear model predicting proteomic levels in pandemic assessment session with additional terms modelling covariates APOE variant status, smoking status, Hip/Waist ratio and Diabetes. Beta estimates have been standardised. NfL – neurofilament light; GFAP (**) indicates significant, FDR corrected by column, alpha=0.05.

Supplementary table 11 Parameter fits for linear model predicting proteomic levels in pandemic assessment session with additional term modelling potential confound Glomerular Filtration Rate (GFR). GFR was available in around half of participants. Beta estimates have been standardised. NfL – neurofilament light; GFAP (**) indicates significant, FDR corrected by column, alpha=0.05.

Supplementary table 12 Parameter fits for interaction term with case-control status (case=+1) and associations between UK Biobank variables at baseline with protein biomarker change. These terms identify UK Biobank variables associated with the protein biomarkers in SARS-CoV-2 exposure specific manner. (*) p<0.05 uncorrected (**) p<0.05 FDR correction (alpha = 0.05).

Supplementary table 13 AD protein level change model fits for models including terms modelling AD neuroimaging phenotype at baseline. Associations were determined from linear models including age and sex covariates. (*) p<0.05 uncorrected (**) p<0.05 FDR correction (alpha = 0.05).

**Supplementary table 14.**
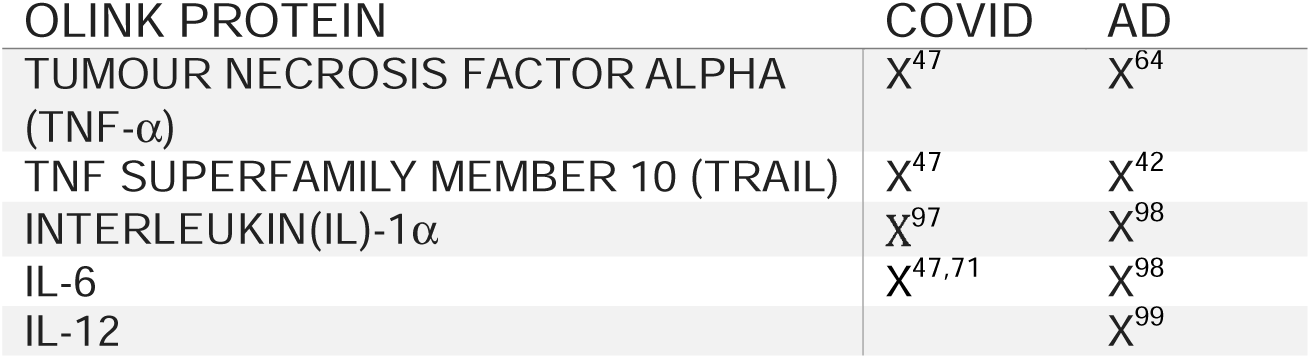
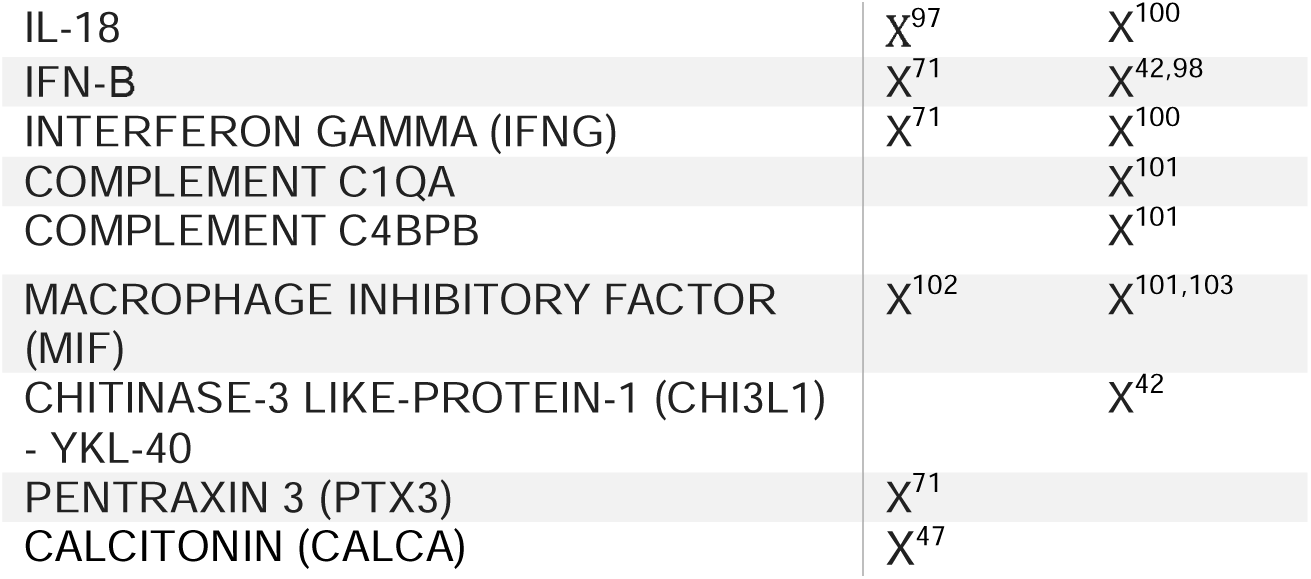
Inflammatory proteins available from Olink panel identified as associated with COVID and/or Alzheimer’s Disease. X indicates reports of an association with the disease.

Supplementary table 15 Baseline Case-Control differences in Olink Inflammatory Proteins. P-values reflect paired t-tests.

Supplementary table 16 Associations of Olink Inflammatory Protein levels (measured at baseline) with the change in AD protein biomarkers across assessments (across cases and controls). Associations were determined from linear models including terms for age, sex, and interval between assessments. (*) p<0.05 uncorrected (**) p<0.05 FDR correction (alpha = 0.05).

Supplementary table 17 Associations of an interaction term between Case/Control status and Olink Inflammatory Protein levels (measured at baseline) with protein biomarker change. These identify Olink Inflammatory Proteins associated with the AD protein biomarkers in a SARS-CoV-2 exposure specific manner. Associations were determined from linear models including terms for age, sex, and interval between assessments. (*) p<0.05 uncorrected (**) p<0.05 FDR correction (alpha = 0.05).

Supplementary table 18 Parameter fits for linear model predicting Olink Inflammatory Protein levels in pandemic assessment session. Beta estimates have been standardised. (**) indicates significant, FDR corrected by column, alpha=0.05.

Supplementary table 19 SARS-CoV-2 related increases in Olink ProteinScore disease risk scores. Plots show SARS-CoV-2 effect weighting parameter in model of change of scores between assessment visits. Due to the ProteinScore disease risk training data, results are from participants <73 years. (*) reflects FDR corrected significance, alpha=0.05.

Supplementary table 20 Correlations between Simoa Ultrasensitive neurology assays. Diagonal reflects correlations of protein levels between pre-pandemic and pandemic assessment visits.

## References

1. Levine, K. S. et al. Virus exposure and neurodegenerative disease risk across national biobanks. Neuron 111, 1086–1093.e2 (2023).

2. Ou, Y.-N. et al. Associations of Infectious Agents with Alzheimer’s Disease: A Systematic Review and Meta-Analysis. Journal of Alzheimer’s Disease 75, 299–309 (2020).

3. Muzambi, R. et al. Assessment of common infections and incident dementia using UK primary and secondary care data: a historical cohort study. The Lancet Healthy Longevity 2, e426–e435 (2021).

4. Muzambi, R. et al. Are infections associated with cognitive decline and neuroimaging outcomes? A historical cohort study using data from the UK Biobank study linked to electronic health records. Transl Psychiatry 12, 1–11 (2022).

5. Debisarun, P. A. et al. Induction of trained immunity by influenza vaccination - impact on COVID-19. PLOS Pathogens 17, e1009928 (2021).

6. Franceschi, C. & Campisi, J. Chronic Inflammation (Inflammaging) and Its Potential Contribution to Age-Associated Diseases. The Journals of Gerontology: Series A 69, S4–S9 (2014).

7. Tejera, D. et al. Systemic inflammation impairs microglial Aβ clearance through NLRP3 inflammasome. The EMBO Journal 38, e101064 (2019).

8. Hosseini, S., Michaelsen-Preusse, K., Schughart, K. & Korte, M. Long-Term Consequence of Non-neurotropic H3N2 Influenza A Virus Infection for the Progression of Alzheimer’s Disease Symptoms. Frontiers in Cellular Neuroscience 15, (2021).

9. Xu, E., Xie, Y. & Al-Aly, Z. Long-term neurologic outcomes of COVID-19. Nat Med 28, 2406–2415 (2022).

10. Douaud, G. et al. SARS-CoV-2 is associated with changes in brain structure in UK Biobank. Nature 604, 697–707 (2022).

11. Hampshire, A. et al. Cognitive deficits in people who have recovered from COVID-19. eClinicalMedicine 39, (2021).

12. Hampshire, A. et al. Cognition and Memory after Covid-19 in a Large Community Sample. N Engl J Med 390, 806–818 (2024).

13. Shan, D., Wang, C., Crawford, T. & Holland, C. Temporal Association between COVID- 19 Infection and Subsequent New-Onset Dementia in Older Adults: A Systematic Review and Meta-Analysis. SSRN Scholarly Paper at 10.2139/ssrn.4716751 (2024).

14. Xie, J. et al. Low-grade peripheral inflammation affects brain pathology in the AppNL-G- Fmouse model of Alzheimer’s disease. acta neuropathol commun 9, 163 (2021).

15. Yang, A. C. et al. Dysregulation of brain and choroid plexus cell types in severe COVID-19. Nature 595, 565–571 (2021).

16. Taquet, M., Geddes, J. R., Husain, M., Luciano, S. & Harrison, P. J. 6-month neurological and psychiatric outcomes in 236 379 survivors of COVID-19: a retrospective cohort study using electronic health records. The Lancet Psychiatry 8, 416–427 (2021).

17. Zetterberg, H. & Blennow, K. Moving fluid biomarkers for Alzheimer’s disease from research tools to routine clinical diagnostics. Molecular Neurodegeneration 16, 10 (2021).

18. Saunders, T. S. et al. Predictive blood biomarkers and brain changes associated with age-related cognitive decline. Brain Communications 5, fcad113 (2023).

19. Wang, L., et al. Plasma proteomics of SARS-CoV-2 infection and severity reveals impact on Alzheimer’s and coronary disease pathways. iScience 26, (2023).

20. Gadd, D. A. et al. Blood protein levels predict leading incident diseases and mortality in UK Biobank. 2023.05.01.23288879 Preprint at 10.1101/2023.05.01.23288879 (2023).

21. Carrasco-Zanini, J. et al. Proteomic prediction of common and rare diseases. 2023.07.18.23292811 Preprint at 10.1101/2023.07.18.23292811 (2023).

22. You, J. et al. Plasma proteomic profiles predict individual future health risk. Nat Commun 14, 7817 (2023).

23. Oh, H. S.-H. et al. Organ aging signatures in the plasma proteome track health and disease. Nature 624, 164–172 (2023).

24. Jia, J. et al. Biomarker Changes during 20 Years Preceding Alzheimer’s Disease. New England Journal of Medicine 390, 712–722 (2024).

25. Janelidze, S. et al. Plasma P-tau181 in Alzheimer’s disease: relationship to other biomarkers, differential diagnosis, neuropathology and longitudinal progression to Alzheimer’s dementia. Nat Med 26, 379–386 (2020).

26. Baldeiras, I. et al. Addition of the Aβ42/40 ratio to the cerebrospinal fluid biomarker profile increases the predictive value for underlying Alzheimer’s disease dementia in mild cognitive impairment. Alzheimer’s Research & Therapy 10, 33 (2018).

27. Fandos, N. et al. Plasma amyloid β 42/40 ratios as biomarkers for amyloid β cerebral deposition in cognitively normal individuals. Alzheimers Dement (Amst) 8, 179–187 (2017).

28. Palmqvist, S. et al. Cerebrospinal fluid and plasma biomarker trajectories with increasing amyloid deposition in Alzheimer’s disease. EMBO Molecular Medicine 11, e11170 (2019).

29. Mattsson, N., Andreasson, U., Zetterberg, H., Blennow, K., & for the Alzheimer’s Disease Neuroimaging Initiative. Association of Plasma Neurofilament Light With Neurodegeneration in Patients With Alzheimer Disease. JAMA Neurology 74, 557–566 (2017).

30. Kim, K. Y., Shin, K. Y. & Chang, K.-A. GFAP as a Potential Biomarker for Alzheimer’s Disease: A Systematic Review and Meta-Analysis. Cells 12, 1309 (2023).

31. Lai, Y.-J. et al. Biomarkers in long COVID-19: A systematic review. Frontiers in Medicine 10, (2023).

32. Frontera, J. A. et al. Comparison of serum neurodegenerative biomarkers among hospitalized COVID-19 patients versus non-COVID subjects with normal cognition, mild cognitive impairment, or Alzheimer’s dementia. Alzheimers Dement 18, 899–910 (2022).

33. Williams, S. A. et al. Plasma protein patterns as comprehensive indicators of health. Nat Med 25, 1851–1857 (2019).

34. Lindbohm, J. V. et al. Plasma proteins, cognitive decline, and 20-year risk of dementia in the Whitehall II and Atherosclerosis Risk in Communities studies. Alzheimer’s & Dementia 18, 612–624 (2022).

35. Sun, B. B. et al. Plasma proteomic associations with genetics and health in the UK Biobank. Nature 622, 329–338 (2023).

36. Schindler, S. E. et al. High-precision plasma β-amyloid 42/40 predicts current and future brain amyloidosis. Neurology 93, e1647–e1659 (2019).

37. Levin, A. T. et al. Assessing the age specificity of infection fatality rates for COVID-19: systematic review, meta-analysis, and public policy implications. European Journal of Epidemiology 35, 1123 (2020).

38. Stocker, H. et al. Association of Kidney Function With Development of Alzheimer Disease and Other Dementias and Dementia-Related Blood Biomarkers. JAMA Network Open 6, e2252387 (2023).

39. Gallo, G., Calvez, V. & Savoia, C. Hypertension and COVID-19: Current Evidence and Perspectives. High Blood Press Cardiovasc Prev 29, 115–123 (2022).

40. Biskaduros, A. et al. Longitudinal trajectories of Alzheimer’s disease CSF biomarkers and blood pressure in cognitively healthy subjects. Alzheimer’s & Dementia n/a,.

41. Camerlingo, C. Vaccination to reduce severe COVID-19 and mortality in COVID-19 patients: a systematic review and meta-analysis. European Review https://www.europeanreview.org/article/28248 (2022).

42. Safdari Lord, J., Soltani Rezaiezadeh, J., Yekaninejad, M. S. & Izadi, P. The association of APOE genotype with COVID-19 disease severity. Sci Rep 12, 13483 (2022).

43. Azevedo, T. et al. Identifying healthy individuals with Alzheimer neuroimaging phenotypes in the UK Biobank. 2022.01.05.22268795 Preprint at 10.1101/2022.01.05.22268795 (2022).

44. Klyucherev, T. O. et al. Advances in the development of new biomarkers for Alzheimer’s disease. Transl Neurodegener 11, 25 (2022).

45. Vergallo, A. et al. Association of plasma YKL-40 with brain amyloid-β levels, memory performance, and sex in subjective memory complainers. Neurobiology of Aging 96, 22– 32 (2020).

46. Lennol, M. P. et al. Transient Changes in the Plasma of Astrocytic and Neuronal Injury Biomarkers in COVID-19 Patients without Neurological Syndromes. Int J Mol Sci 24, 2715 (2023).

47. Byeon, S. K. et al. Development of a multiomics model for identification of predictive biomarkers for COVID-19 severity: a retrospective cohort study. Lancet Digit Health 4, e632–e645 (2022).

48. Gaudet, L. A. et al. Associations between SARS-CoV-2 infection and incidence of new chronic condition diagnoses: a systematic review. Emerg Microbes Infect 12, 2204166.

49. Fawns-Ritchie, C. & Deary, I. J. Reliability and validity of the UK Biobank cognitive tests. PLOS ONE 15, e0231627 (2020).

50. Atchison, C. J. et al. Long-term health impacts of COVID-19 among 242,712 adults in England. Nat Commun 14, 6588 (2023).

51. Mizrahi, B. et al. Long covid outcomes at one year after mild SARS-CoV-2 infection: nationwide cohort study. BMJ 380, e072529 (2023).

52. Boulo, S. et al. First amyloid β1-42 certified reference material for re-calibrating commercial immunoassays. Alzheimer’s & Dementia 16, 1493–1503 (2020).

53. Virhammar, J. et al. Biomarkers for central nervous system injury in cerebrospinal fluid are elevated in COVID-19 and associated with neurological symptoms and disease severity. European Journal of Neurology 28, 3324–3331 (2021).

54. Therriault, J. et al. Association of Phosphorylated Tau Biomarkers With Amyloid Positron Emission Tomography vs Tau Positron Emission Tomography. JAMA Neurol 80, 188– 199 (2023).

55. van Arendonk, J. et al. Plasma neurofilament light chain in relation to 10-year change in cognition and neuroimaging markers: a population-based study. GeroScience (2023) doi:10.1007/s11357-023-00876-5.

56. O’Connor, A. et al. Plasma GFAP in presymptomatic and symptomatic familial Alzheimer’s disease: a longitudinal cohort study. J Neurol Neurosurg Psychiatry 94, 90– 92 (2023).

57. Diray-Arce, J., et al. Multi-omic longitudinal study reveals immune correlates of clinical course among hospitalized COVID-19 patients. CR Med 4, (2023).

58. Zhang, X., Fu, Z., Meng, L., He, M. & Zhang, Z. The Early Events That Initiate β-Amyloid Aggregation in Alzheimer’s Disease. Front Aging Neurosci 10, 359 (2018).

59. Abbott, A. Are infections seeding some cases of Alzheimer’s disease? Nature 587, 22– 25 (2020).

60. Mawanda, F., Wallace, R. B., McCoy, K. & Abrams, T. E. Systemic and localized extra- central nervous system bacterial infections and the risk of dementia among US veterans: A retrospective cohort study. *Alzheimer’s & Dementia: Diagnosis*, Assessment & Disease Monitoring 4, 109–117 (2016).

61. Sipilä, P. N. et al. Hospital-treated infectious diseases and the risk of dementia: a large, multicohort, observational study with a replication cohort. The Lancet Infectious Diseases 21, 1557–1567 (2021).

62. Radke, J. et al. Proteomic and transcriptomic profiling of brainstem, cerebellum and olfactory tissues in early- and late-phase COVID-19. Nat Neurosci 27, 409–420 (2024).

63. Gordon, M. N. et al. Impact of COVID-19 on the Onset and Progression of Alzheimer’s Disease and Related Dementias: A Roadmap for Future Research. Alzheimer’s & Dementia 18, 1038–1046 (2022).

64. Walker, K. A. et al. The role of peripheral inflammatory insults in Alzheimer’s disease: a review and research roadmap. Molecular Neurodegeneration 18, 37 (2023).

65. Jorfi, M., Maaser-Hecker, A. & Tanzi, R. E. The neuroimmune axis of Alzheimer’s disease. Genome Med 15, 6 (2023).

66. Holmes, C. et al. Systemic inflammation and disease progression in Alzheimer disease. Neurology 73, 768–774 (2009).

67. Morgan, D. G. & Mielke, M. M. Knowledge gaps in Alzheimer’s disease immune biomarker research. Alzheimer’s & Dementia 17, 2030–2042 (2021).

68. Kitazawa, M., Oddo, S., Yamasaki, T. R., Green, K. N. & LaFerla, F. M. Lipopolysaccharide-Induced Inflammation Exacerbates Tau Pathology by a Cyclin- Dependent Kinase 5-Mediated Pathway in a Transgenic Model of Alzheimer’s Disease. J. Neurosci. 25, 8843–8853 (2005).

69. Walker, K. A. et al. Midlife systemic inflammatory markers are associated with late-life brain volume: The ARIC study. Neurology 89, 2262 (2017).

70. Jacob, F. et al. Human Pluripotent Stem Cell-Derived Neural Cells and Brain Organoids Reveal SARS-CoV-2 Neurotropism Predominates in Choroid Plexus Epithelium. Cell Stem Cell 27, 937–950.e9 (2020).

71. Phetsouphanh, C. et al. Immunological dysfunction persists for 8 months following initial mild-to-moderate SARS-CoV-2 infection. Nat Immunol 23, 210–216 (2022).

72. Hur, J.-Y. et al. The innate immunity protein IFITM3 modulates γ-secretase in Alzheimer’s disease. Nature 586, 735–740 (2020).

73. Gholami, M. et al. Interferon-Induced Transmembrane Protein 3 rs34481144 C/T Genotype and Clinical Parameters Related to Progression of COVID-19. J Immunol Res 2023, 2345062 (2023).

74. Ashton, N. J. et al. Diagnostic Accuracy of a Plasma Phosphorylated Tau 217 Immunoassay for Alzheimer Disease Pathology. JAMA Neurol 81, 255–263 (2024).

75. Janelidze, S. et al. Plasma β-amyloid in Alzheimer’s disease and vascular disease. Sci Rep 6, 26801 (2016).

76. Xie, Y., Xu, E., Bowe, B. & Al-Aly, Z. Long-term cardiovascular outcomes of COVID-19. Nat Med 28, 583–590 (2022).

77. Wan, E. Y. F. et al. Association of COVID-19 with short- and long-term risk of cardiovascular disease and mortality: a prospective cohort in UK Biobank. Cardiovascular Research 119, 1718–1727 (2023).

78. Marín, J. S. et al. Increased incidence of rheumatoid arthritis after COVID-19. Autoimmunity Reviews 22, 103409 (2023).

79. Rathmann, W., Kuss, O. & Kostev, K. Incidence of newly diagnosed diabetes after Covid-19. Diabetologia 65, 949–954 (2022).

80. Argentieri, M. A. et al. Proteomic aging clock predicts mortality and risk of common age- related diseases in diverse populations. 2023.09.13.23295486 Preprint at 10.1101/2023.09.13.23295486 (2023).

81. Livingston, G. et al. Dementia prevention, intervention, and care: 2020 report of the Lancet Commission. The Lancet 396, 413–446 (2020).

82. Wang, Y.-R. et al. Associations Between Hepatic Functions and Plasma Amyloid-Beta Levels-Implications for the Capacity of Liver in Peripheral Amyloid-Beta Clearance. Mol Neurobiol 54, 2338–2344 (2017).

83. Lyu, H. et al. Plasma amyloid-beta levels correlated with impaired hepatic functions: An adjuvant biomarker for the diagnosis of biliary atresia. Front. Surg. 9, (2022).

84. Cousins, K. A. Q. et al. Elevated Plasma Phosphorylated Tau 181 in Amyotrophic Lateral Sclerosis. Annals of Neurology 92, 807–818 (2022).

85. Vacchiano, V. et al. Elevated plasma p-tau181 levels unrelated to Alzheimer’s disease pathology in amyotrophic lateral sclerosis. J Neurol Neurosurg Psychiatry 94, 428–435 (2023).

86. Schoeler, T. et al. Participation bias in the UK Biobank distorts genetic associations and downstream analyses. Nat Hum Behav 7, 1216–1227 (2023).

87. Janelidze, S. et al. Head-to-head comparison of 10 plasma phospho-tau assays in prodromal Alzheimer’s disease. Brain 146, 1592–1601 (2022).

88. Ashton, N. J., et al. Plasma and CSF biomarkers in a memory clinic: Head-to-head comparison of phosphorylated tau immunoassays. Alzheimer’s & Dementia 19, 1913–1924 (2023).

89. Wang, L. et al. Association of COVID-19 with New-Onset Alzheimer’s Disease. J Alzheimers Dis 89, 411–414 (2022).

90. Littlejohns, T. J. et al. The UK Biobank imaging enhancement of 100,000 participants: rationale, data collection, management and future directions. Nature Communications 11, 2624 (2020).

91. Stevenson-Hoare, J. et al. Plasma biomarkers and genetics in the diagnosis and prediction of Alzheimer’s disease. Brain 146, 690–699 (2023).

92. Inker Lesley A., et al. Estimating Glomerular Filtration Rate from Serum Creatinine and Cystatin C. New England Journal of Medicine 367, 20–29 (2012).

93. Bešević, J. et al. Persistence of SARS-CoV-2 antibodies over 18 months following infection: UK Biobank COVID-19 Serology Study. J Epidemiol Community Health (2023) doi:10.1136/jech-2023-220569.

94. Second-generation PLINK: rising to the challenge of larger and richer datasets | GigaScience | Oxford Academic. https://academic.oup.com/gigascience/article/4/1/s13742-015-0047-8/2707533?login=false.

95. Miller, K. L. et al. Multimodal population brain imaging in the UK Biobank prospective epidemiological study. Nature Neuroscience 19, 1523–1536 (2016).

96. Alfaro-Almagro, F. et al. Image processing and Quality Control for the first 10,000 brain imaging datasets from UK Biobank. NeuroImage 166, 400–424 (2018).

97. Makaremi, S. et al. The role of IL-1 family of cytokines and receptors in pathogenesis of COVID-19. Inflamm. Res. 71, 923–947 (2022).

98. Dursun, E. et al. The interleukin 1 alpha, interleukin 1 beta, interleukin 6 and alpha-2- macroglobulin serum levels in patients with early or late onset Alzheimer’s disease, mild cognitive impairment or Parkinson’s disease. Journal of Neuroimmunology 283, 50–57 (2015).

99. Yang, H.-S. et al. Plasma IL-12/IFN-γ axis predicts cognitive trajectories in cognitively unimpaired older adults. Alzheimers Dement 18, 645–653 (2022).

100. Sutinen, E. M., Pirttilä, T., Anderson, G., Salminen, A. & Ojala, J. O. Pro-inflammatory interleukin-18 increases Alzheimer’s disease-associated amyloid-β production in human neuron-like cells. Journal of Neuroinflammation 9, 199 (2012).

101. Brosseron, F. et al. Soluble TAM receptors sAXL and sTyro3 predict structural and functional protection in Alzheimer’s disease. Neuron 110, 1009–1022.e4 (2022).

102. Shin, J. J. et al. MIF is a common genetic determinant of COVID-19 symptomatic infection and severity. QJM 116, 205–212 (2023).

103. Nasiri, E. et al. Key role of MIF-related neuroinflammation in neurodegeneration and cognitive impairment in Alzheimer’s disease. Mol Med 26, 34 (2020).

